# Prognostic value of the level of gut microbe-generated metabolite trimethylamine-N-oxide in patients with heart failure: a meta-analysis and systematic review

**DOI:** 10.1101/2021.05.04.21256606

**Authors:** Miao Li-na, Du Jian-peng, Guo Ming, Chen Zhu-hong, Qu Hua, Shi Da-zhuo

## Abstract

**Aims:** Intestinal microbial metabolite trimethylamine N-oxide (TMAO) is considered to be a key mediator between intestinal microbiota changes and heart failure (HF). Clinical evidences showed that patients with HF had relatively high level of plasma TMAO than those without. However, the relationship between plasma TMAO and the prognostics of patients with HF has not been investigated. This study aims to evaluate the relationship between plasma TMAO and the outcomes of patients with HF.

**Methods and results:** PubMed, EMBASE, Cochrane Library databases were included for systematic literature search. The included studies were used for the extraction of overall hazard ratios of adverse events data (myocardial infarction, cardiovascular hospitalization, cardiovascular mortality, revascularization and stroke) as well as all‐cause mortality data. Cochran’s Q test and I^2^ statistical methods were applied to assess heterogeneity. Selection of random effects model or fixed effects model were according to heterogeneity. Sensitivity analysis and subgroup analysis were used to find the source of heterogeneity. Among the 7 included studies, all studies published all‐cause mortality data and 6 provided adverse events. And the results demonstrated that higher level of plasma TMAO (defined according to original studies)was an indicator of risk of all‐cause mortality (HR =0.26, 95% CI: 0.20-0.32, I^2^ = 96.1%,) and risk of adverse events (HR =1.79, 95% CI: 1.37-2.22, I^2^ = 78.6%). There was no significant publication bias among the included studies (Egger’s,P = 0.130;Begg’s tests P = 0.452). Further subgroup analysis did not show that study location, follow-up time, or year of publication had any effect on the results. The results of sensitivity analysis do not indicate that any of the included affected the robustness of the results.

**Conclusion:** Heart failure patients with higher baseline plasma TMAO is a risk factor for all-cause mortality and adverse events.

## Introduction

Heart failure (HF) is a complex multifactor illness that leads to enormous health and socio-economic burden [1]. In vivo studies using animal models and humans suggested that intestinal microbiota likely played a big role in the development of a variety of diseases, including heart failure [2]. Recent studies demonstrated that HF patients had significantly changes in both intestinal microbiota and intestinal permeability [3]. Further research found that TMAO, a metabolite of intestinal flora, was likely connected to the disease emergence and progression of HF [4]. Some studies have pointed out that TMAO level in patients with heart failure is relatively high compared with patients without and is related to the mortality and prognosis of heart failure [3]. However, the relationship between TMAO and the outcomes of HF has not been evaluated. To investigate the relationship, systematic review and meta-analysis of all existing cohort publications to assess the relationship between plasma TMAO at baseline level and the prognostic outcome in HF patients.

## Methods

### Systematic literature search method

PubMed, EMBASE, and Cochrane library databases were chosen for search engines for relevant studies from the inception up to May 2020, without language restrictions. The methodology followed the PRISMA statement used in a systematic review and Meta-analysis of Observational Studies in Epidemiology (MOOSE) guidelines [5]. Key words used are listed in the following: “Heart Failure” AND “2-Hydroxy-N, N, N-trimethylethanaminium “OR “Choline” OR “Trimethylamine “OR “trimethylamine N-oxide” OR “TMAO” OR “trimethylamine oxide” OR “betaine “.

### Inclusion criteria

The current inclusion criteria for systemic review and meta-analysis were as follows: (1) in vivo clinical trials studies; (2) evaluation of plasma TMAO levels at baseline; (3) Adverse outcome (MI, hospitalization due to cardiovascular incidents, cardiovascular mortality, revascularization and stroke) or all-cause mortality as the clinical endpoint; (4) hazard ratios (HR) with reported confidence intervals (CI). Studies in animal trials, case reports or studies with insufficient pooling data were excluded. The systematic literature review of the literatures was conducted by two independent individual reviewers. When dispute occur, consultation with a senior investigator was followed to resolve discrepancies. This paper has been registered in INPLASY database (registration number: INPLASY202010047).

### Data extraction

Information about the first author’s surname, country of origin, year of publication, study design, gender, sample size, age, follow-up period, disease outcome, plasma TMAO, adjusted variable, HR with 95% CI were collected from each included study. The results of meta-analysis included all-cause mortality and adverse outcomes. The data used were collected by two reviewers using a pre-designed data extraction form. The disagreement was settled by involvement of senior researcher.

### Quality evaluation

Newcastle–Ottawa Scale [6] was applied to assess the risk of bias in included publications based on group comparability, group selection, exposure or outcome determination. The quality of each study was assessed by two individuals separately. Disagreement were settled by consulting a senior investigator.

### Statistical analysis

HRs and their CIs were chosen as measurement with the comparison between the highest quartile and the lowest quartile of TMAO levels. The definition of TMAO level is according to the definition of the studies themselves. The Cochran’s Q test was applied to evaluate the heterogeneity among studies, and I^2^ static variable was applied to quantify heterogeneity. For all meta‐analyses in this study, random‐effects model was applied. To test heterogeneity, country of origin, duration of follow-up and year of publication were chosen factors in further analyses. To test the robustness of this study, we performed sensitivity analysis on each single study. Publication bias and selective reporting were examined using funnel plot for visual presentation, where standard error of logarithm HR was plotted against HR. Publication bias was conducted according to Egger’s and Begg’s linear regression tests [7]. All statistical analyses were performed in STATA 12.0. A two‐sided P value <0.05 was considered with statistical significance.

## Results

### Literature search

In total, there were 272 records from PubMed and EMBASE, Cochrane Library databases after key word searching. After removing 94 duplicates, we identified 178 potential studies from our initial search and excluded 161 studies including reviews, experience summaries, animal model experiments, medical record reports. The full texts of the remaining 17 studies were reviewed. Studies that was not fitting to the inclusion criteria and those that non-conformity in terms of the outcome indices were excluded. Eventually, 7 English-language studies met our inclusion criteria in Figure1, study selection is explained by the flow chart.

**Figure1.**
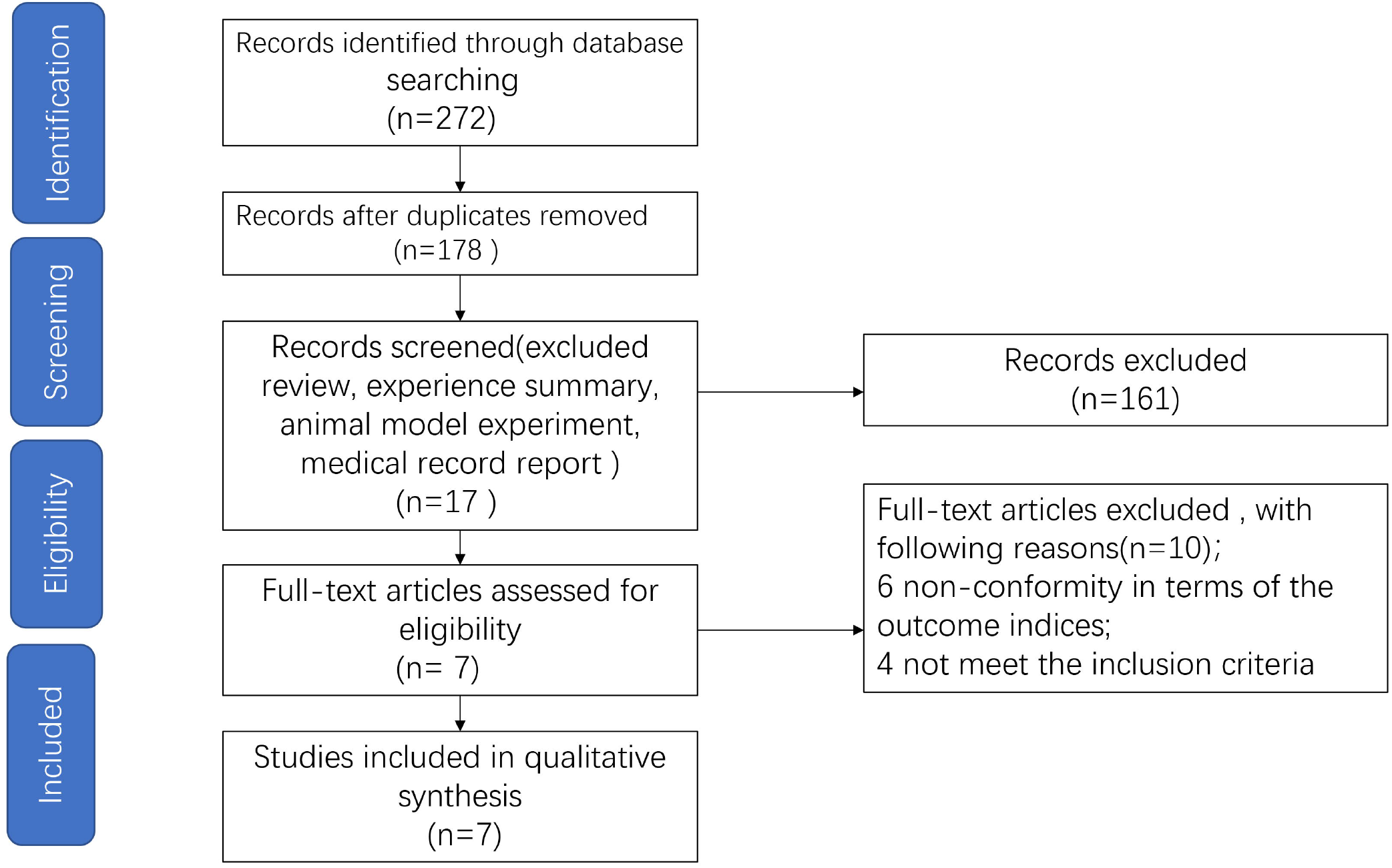
Flow chart of study search and selection.

### Study characteristics

Detailed comparisons of the seven prospective publications between 2014 and 2020 are listed in Table 1. The country of origin of two of these studies were the United States [8; 9], the other two the UK [10; 11]. Others were performed in Norway [12], European [3] and China [13] respectively. The follow-up time of the included studies vary from 1 year to 5.2 years. In general, sample sizes varied from 112 to 2234, and a total of 5,519 samples were included in the meta-analysis in the end. End points extracted from these studies were all-cause mortality and adverse outcomes, including cardiovascular mortality, myocardial infarction, cardiovascular hospitalization, revascularization and stroke. Quality of the studies were of medium to high degree, as shown by the individual NOS scores varying from 6 to 8.

**Table 1.**
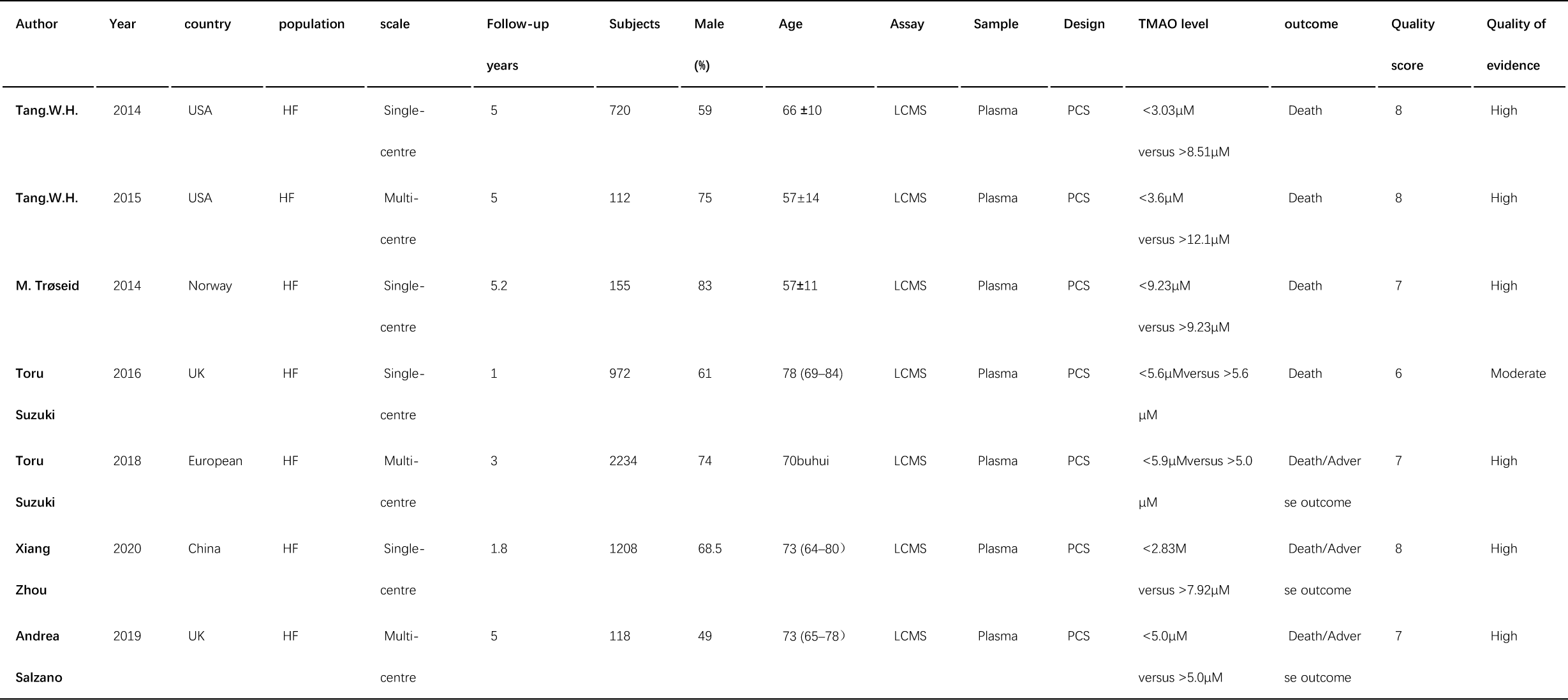
Characteristics of included prospective cohort studies

### TMAO and Adverse Outcomes

Six prospective patient cohorts with a total of 4799 samples presented HRs data for baseline TMAO and adverse outcome risk analyses [3; 9; 10; 11; 12; 13]. The results show that a upward trend of TMAO levels is associated with a upward trend of the risk of adverse outcomes (HR 1.79, 95% CI: 1.37–2.22, P<0.001, I2 = 78.6%). However, a considerably large heterogeneity was observed among various studies I^2^ = 78.6%, P = 0.000; (Figure.2). Therefore, a subgroup analysis was conducted to assess the potential impact of the country of origin, year of follow-up, and publication time of the studies. The discovery was that the pooled HR from studies performed in the United States (HR 1.48, 95% CI: 1.11-1.98) and those in other countries (HR 2.04, 95% CI: 1.48, 2.81, P<0.001, I^2^ = 88.1%) was not statistically significant difference risk of adverse events experienced by patients with increased TMAO levels (Figure3). In addition, subgroup analyses indicated resembling increases of adverse outcome risk in studies with various follow-up time (follow-up <5 years: HR 1.91, 95% CI: 1.31-2.79, P<0.001, I^2^ = 93.2%; follow-up ≥5 years: HR 1.88, 95% CI: 1.29, 2.75, P = 0.172, I^2^ = 43.2%) (Figure4). The association of baseline TMAO and adverse outcome presented resembling increases of adverse outcome risk in studies before 2015 (HR 1.69, 95% CI: 1.16-2.48, P = 0.197, I^2^ = 39.9%) and after 2015 (HR 2.01, 95% CI: 1.41-2.87, P<0.001, I^2^ = 90.6%) (Figure 5) (Table 2). Based on this result, none of the studies were responsible for the heterogeneity observed. Funnel plot did not show obvious asymmetry, suggesting low risk of publication bias. Nevertheless, visual examination of the results of the Egger’s (P = 0.130) and Begg’s tests (P = 0.452) indicated no significant publication bias (figure 6,7). The results of the sensitivity analysis did not indicate that any study affected the robustness of the study (Figure 8).

**Table 2.**
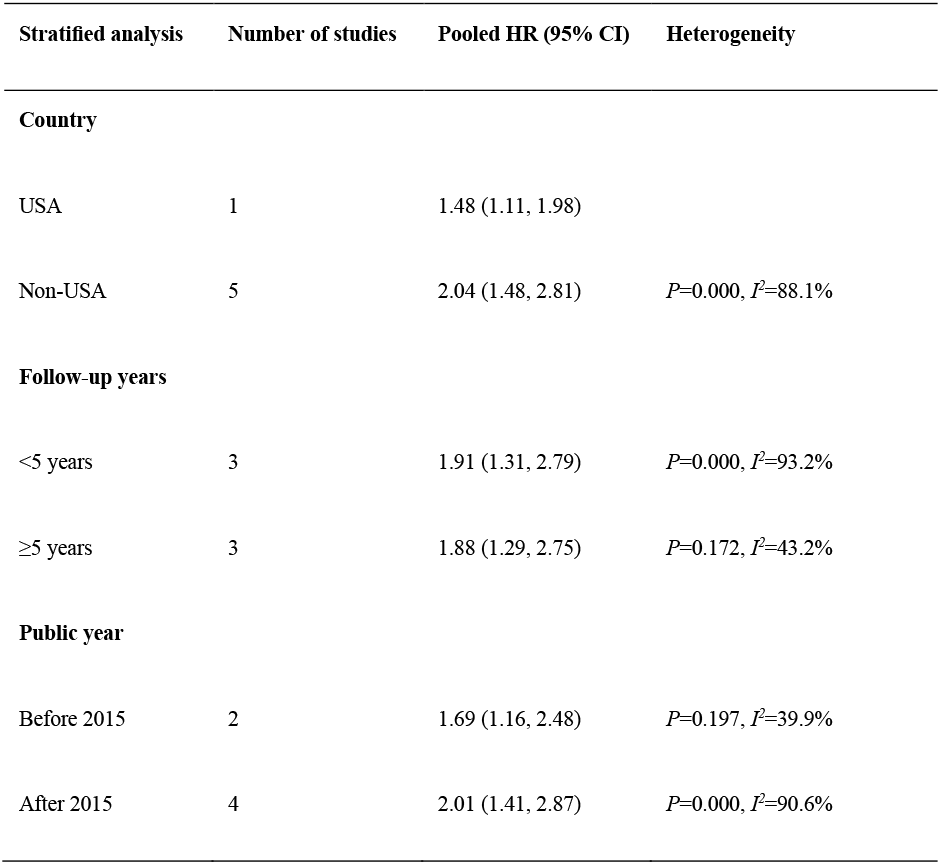
Stratified analyses of pooled hazard risks of TMAO and adverse outcome

**Figure 2.**
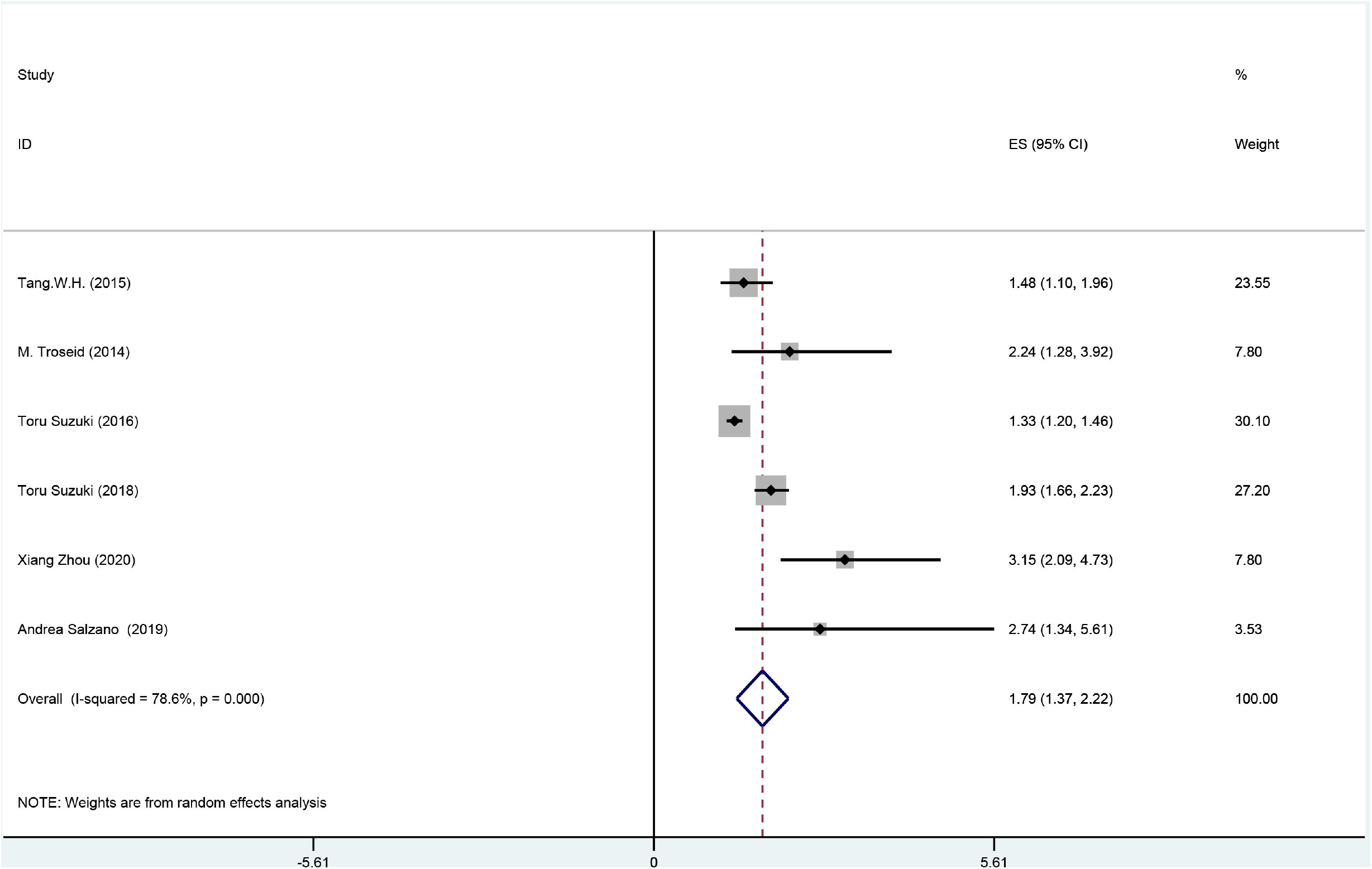
Forest plot for the association between baseline TMAO and adverse outcomes.

**Figure 3.**
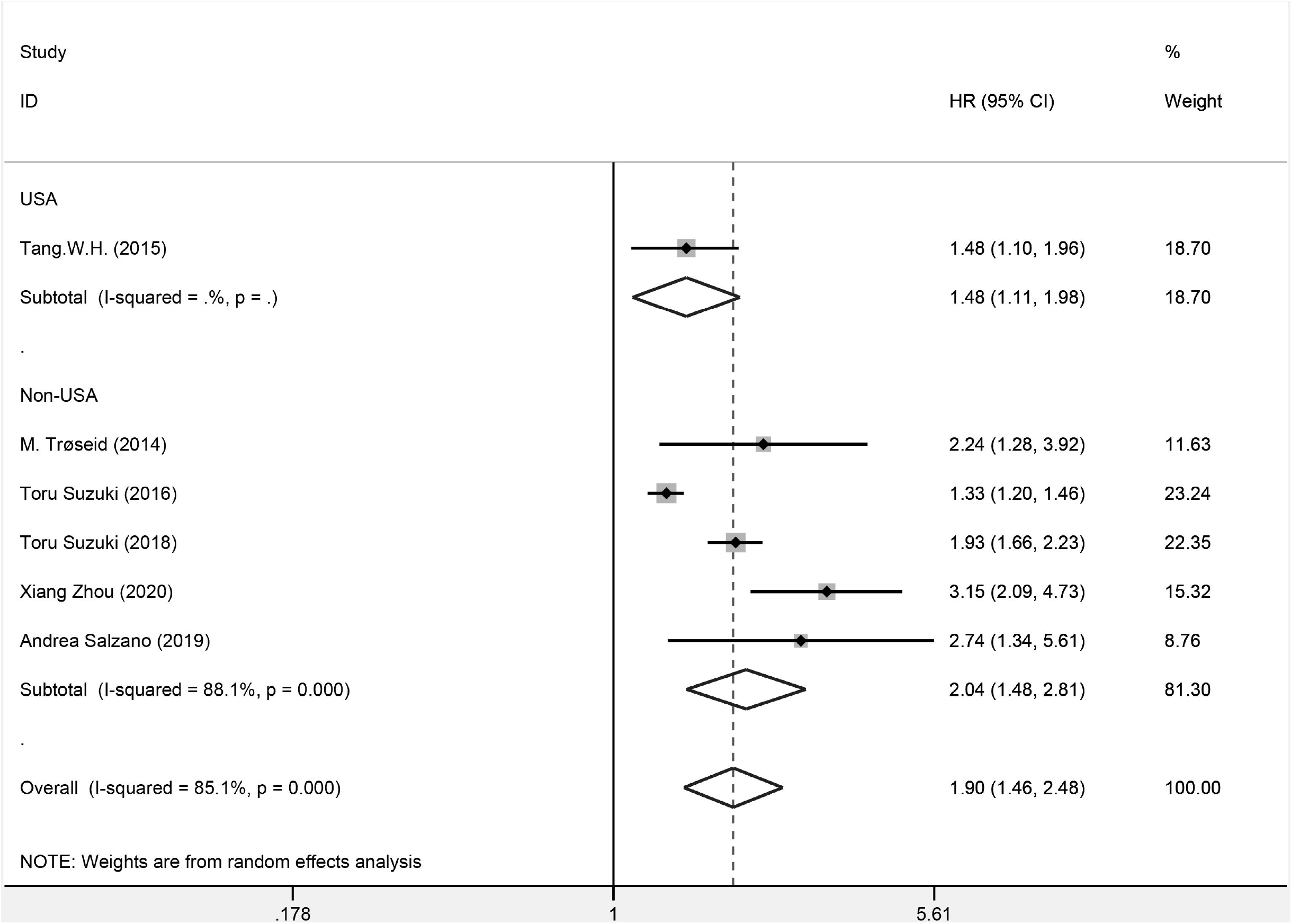
Forest plot for the association between TMAO and adverse outcomes in the United States and those in other countries.

**Figure 4.**
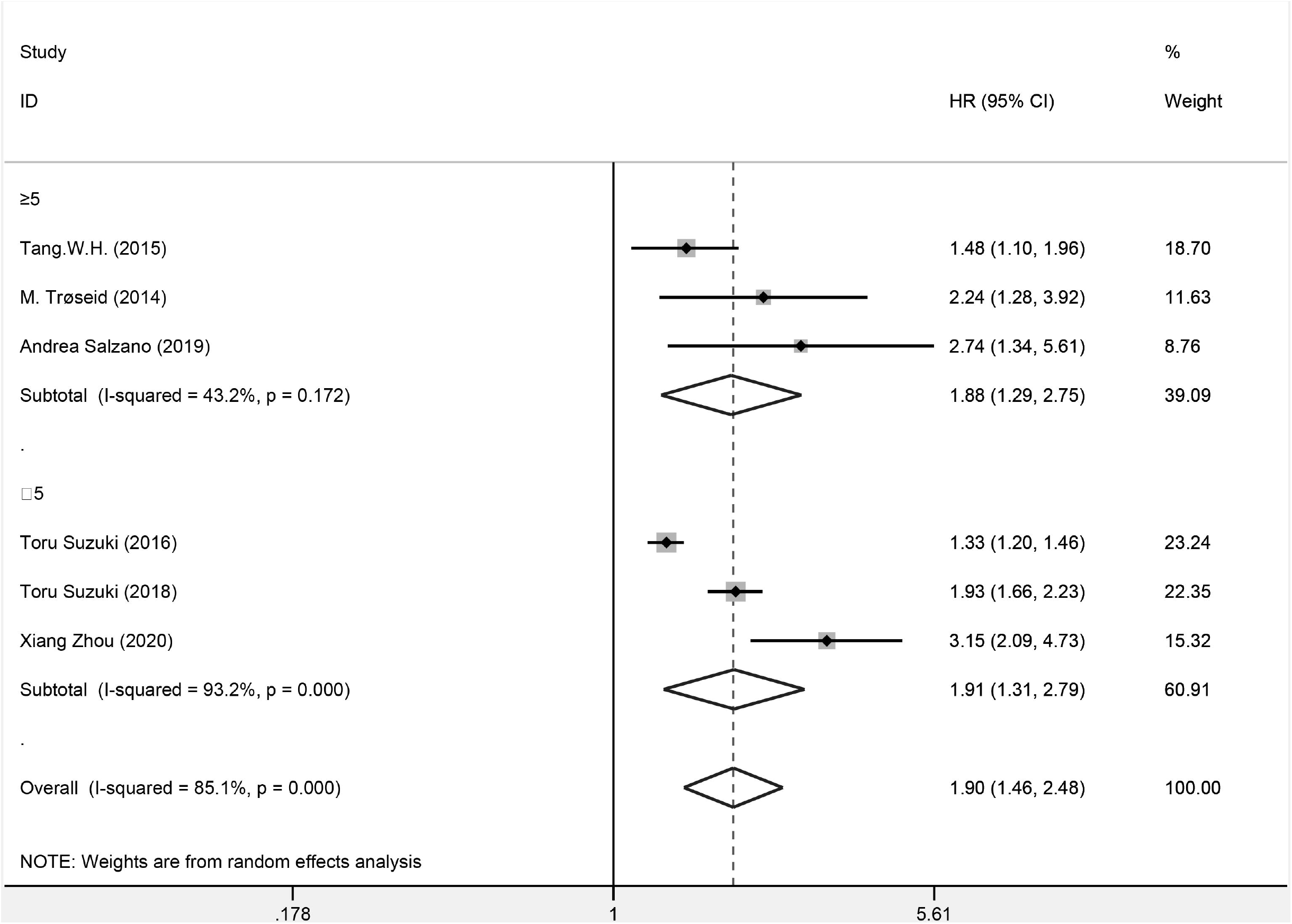
Forest plot for the association between TMAO and adverse outcomes with different follow-up periods.

**Figure 5.**
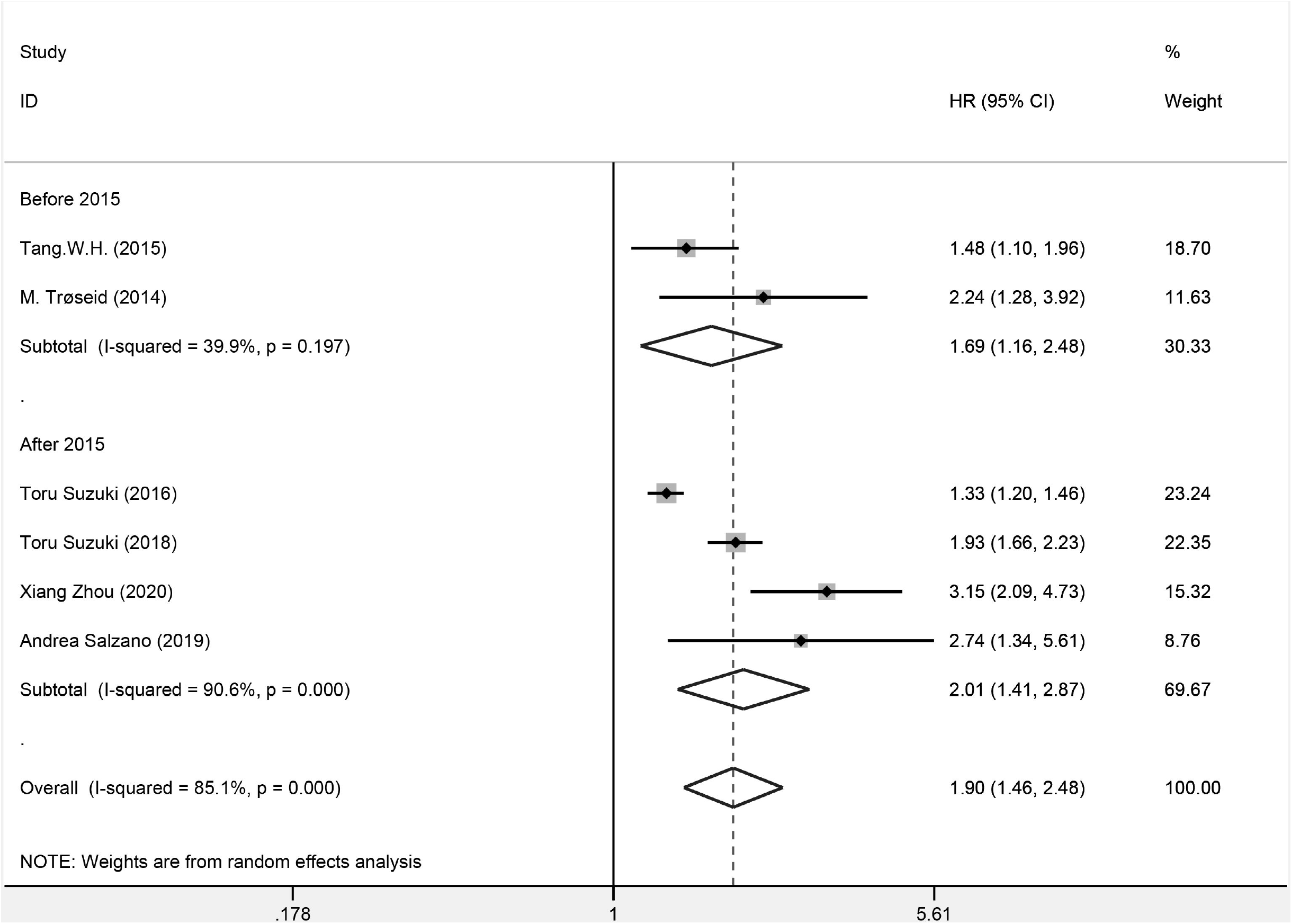
Forest plot for the association between TMAO and adverse outcomes in studies before 2015 and after 2015.

**Figure 6.**
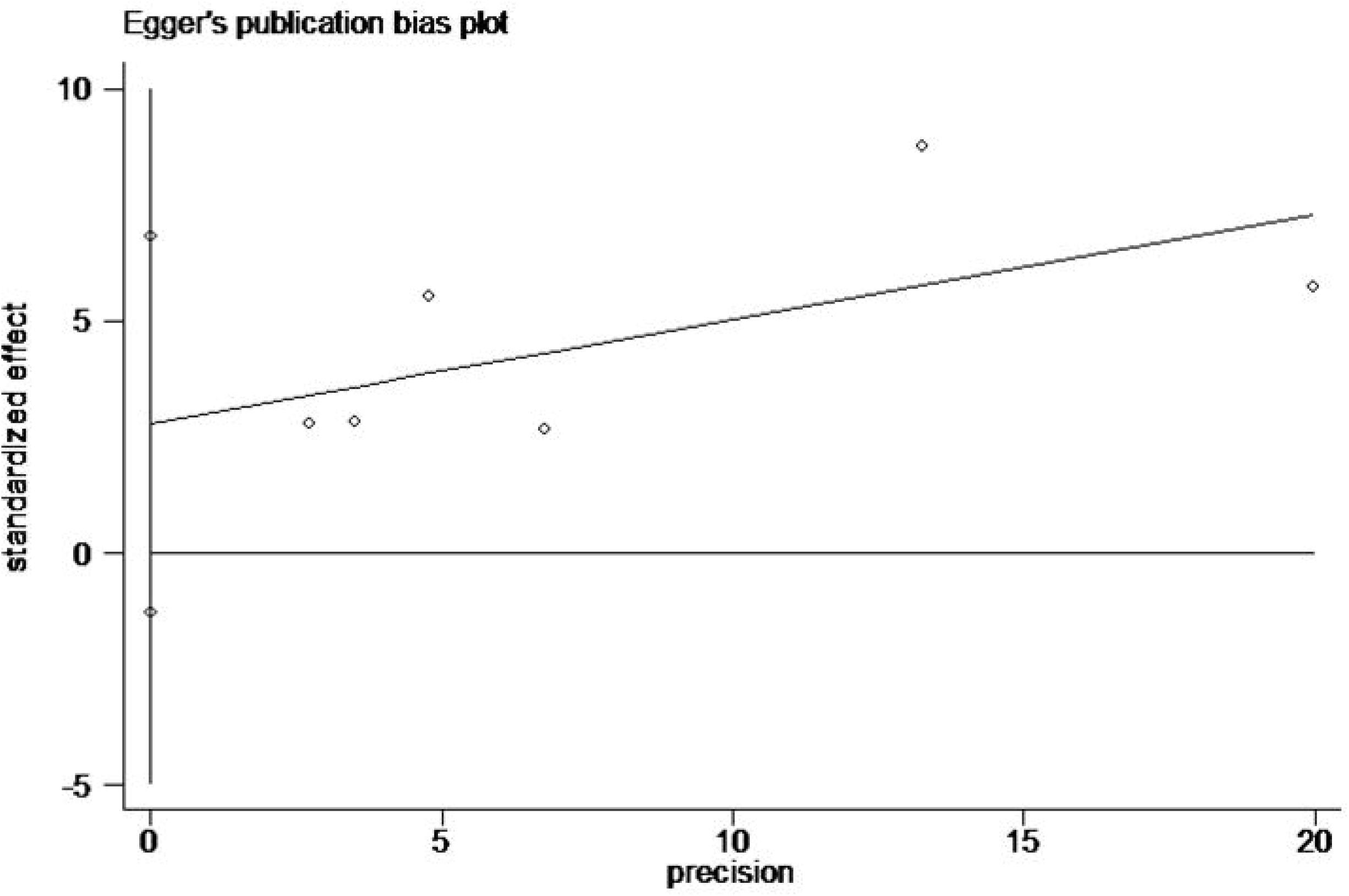
Egger’s publication bias plot for the association between baseline TMAO and adverse outcomes.

**Figure 7.**
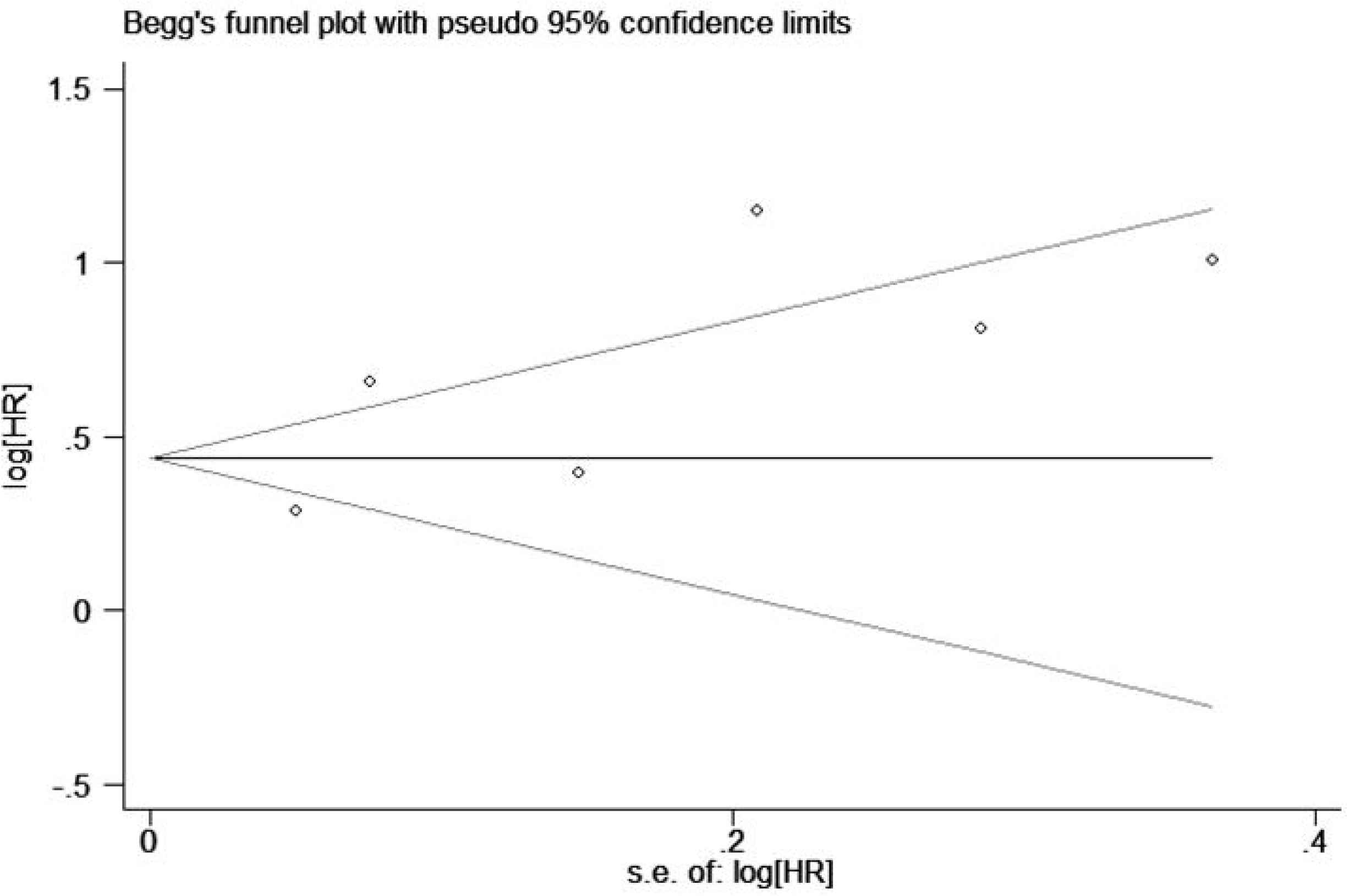
Begg’s publication bias plot for the association between baseline TMAO and adverse outcomes.

**Figure 8.**
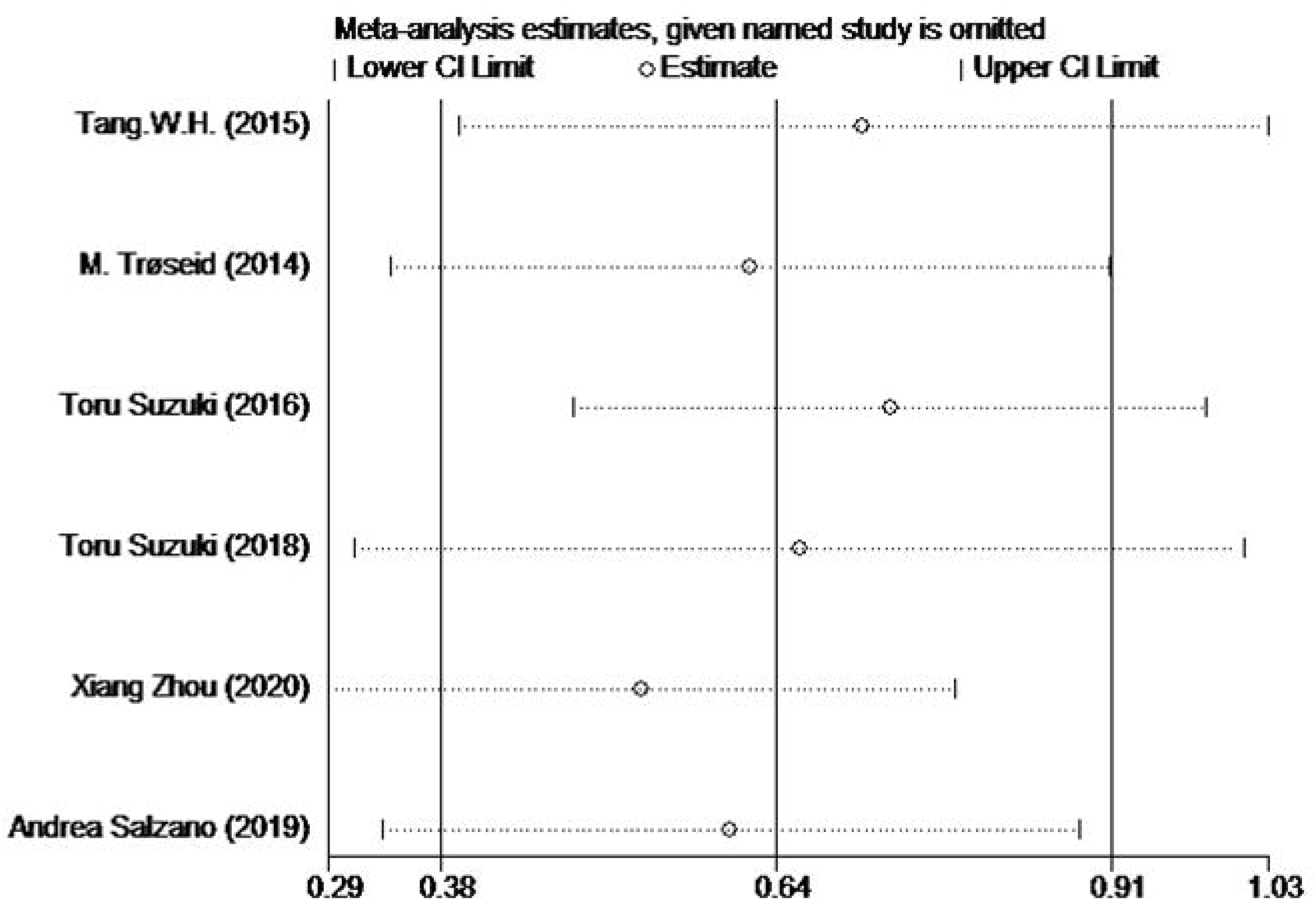
Sensitivity analysis for TMAO level and adverse outcomes.

### TMAO and All‐cause mortality

7 studies with 5519 samples indicated a potential association between baseline TMAO and all-cause mortality [3; 8; 9; 10; 11; 12; 13]. Meta-analysis presented that the upward trend of TMAO levels were independently associated with that of all-cause mortality (HR 0.26, 95% CI: 0.20-0.32, P<0.001 I^2^ = 96.1%). However, a considerably big heterogeneity was discovered among different studies (I^2^ = 96.1%, P<0.001) (Figure.9). Therefore, we performed subgroup analyses to assess the potential impact of the country of origin, follow-up year, and publication time of the studies. It is discovered afterwards that, the pooled HR from the American studies (HR 0.31, 95% CI: 0.25-0.37, P = 0.149 I^2^= 51.9%) and those in other countries (HR 0.24, 95% CI: 0.16, 0.31, P<0.001, I^2^ = 96.9%) was not statistically significant difference risk of all-cause mortality in patients with increased TMAO levels (Figure.10). Additionally, subgroup analyses suggested similar increases in all-cause mortality among studies with different follow-up time (follow-up <5 years: HR 0.22, 95% CI: 0.13-0.32, P<0.001, I^2^= 98.4%; follow-up ≥5 years: HR 0.29, 95% CI: 0.25-0.32, P=0.311, I^2^ = 16.2%)(Figure.11). The correlation of baseline TMAO and all-cause mortality risk still stick to the same conclusion despite the addition of more recent studies (P<0.001) (Table 3). Nonetheless, Egger’s (P = 0.387, Figure13) and Begg’s tests(P = 0.764, Figure14) did not indicate risk of publication bias. Sensitivity did not suggest any study could impact the robustness of the study (Figure15).

**Table 3.**
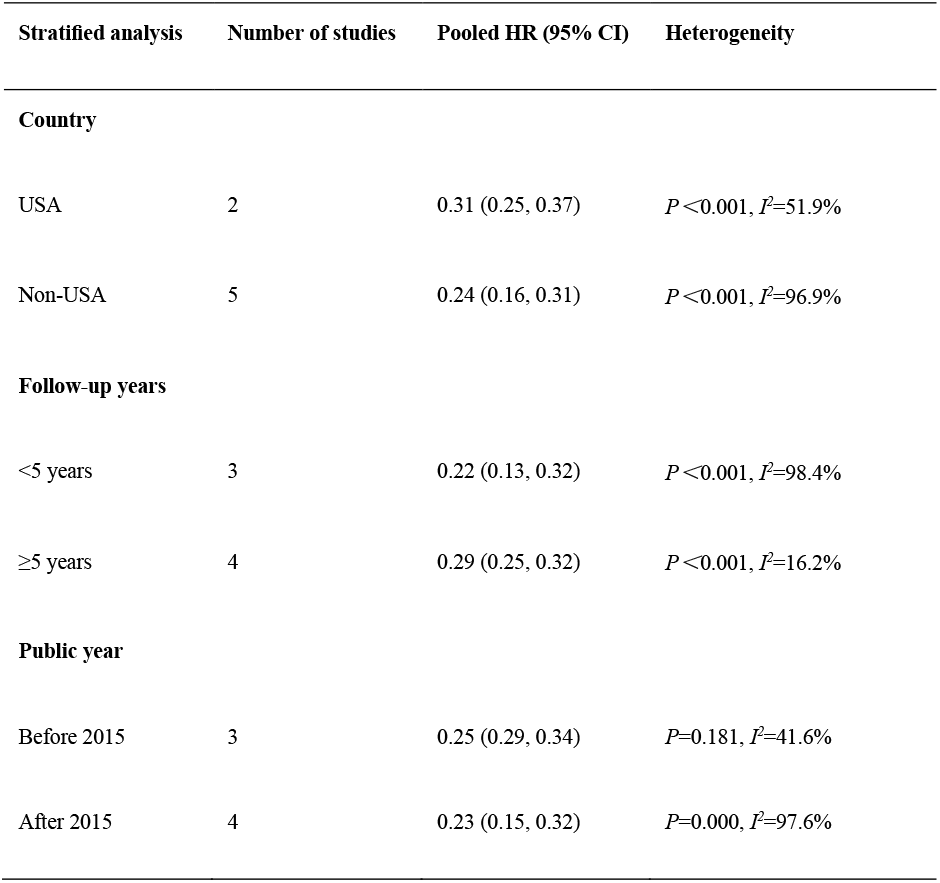
Stratified analyses of pooled hazard risks of TMAO and All‐cause mortality

**Figure 9.**
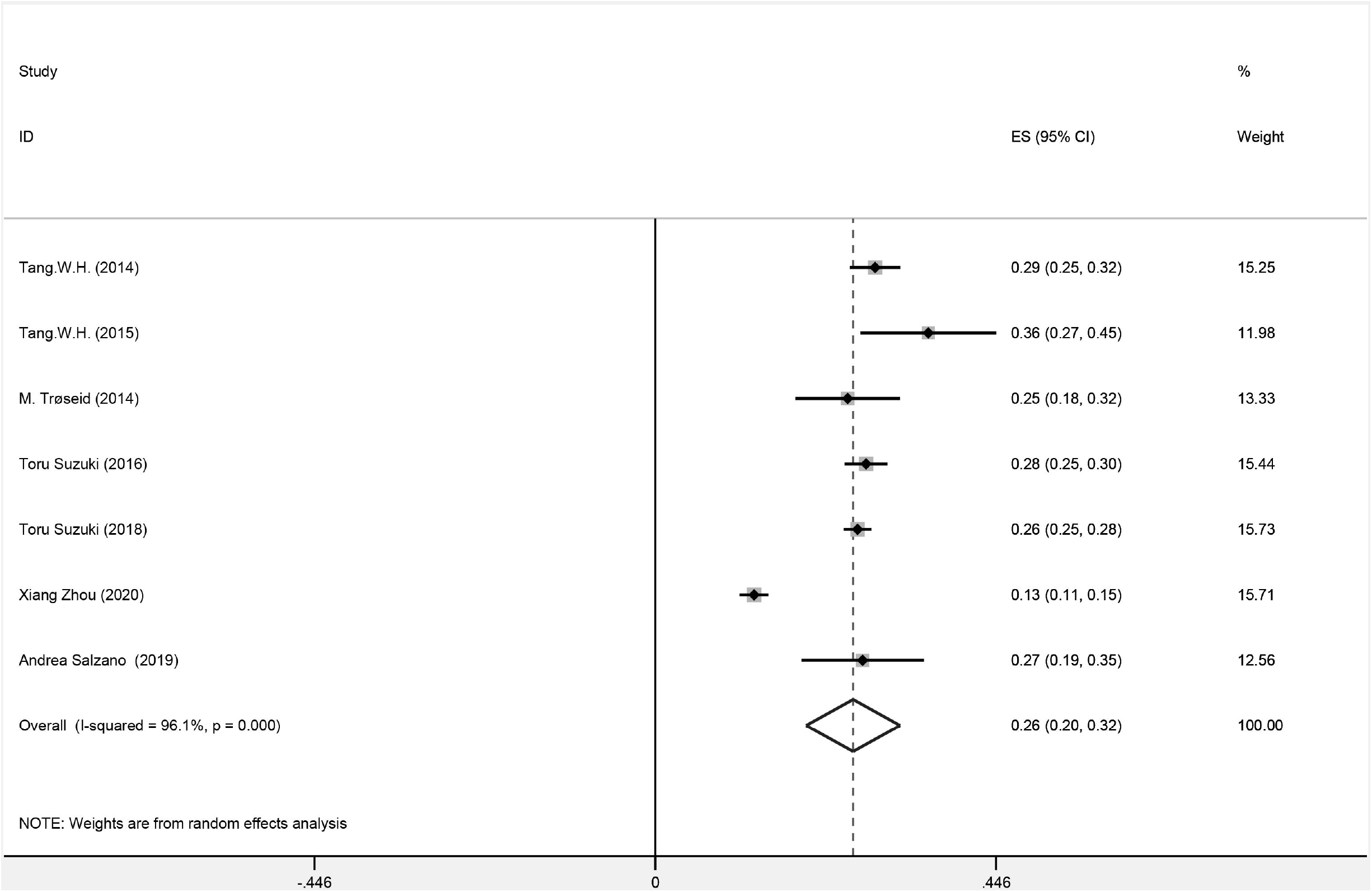
Forest plot for the association between baseline TMAO and All‐cause mortality.

**Figure 10.**
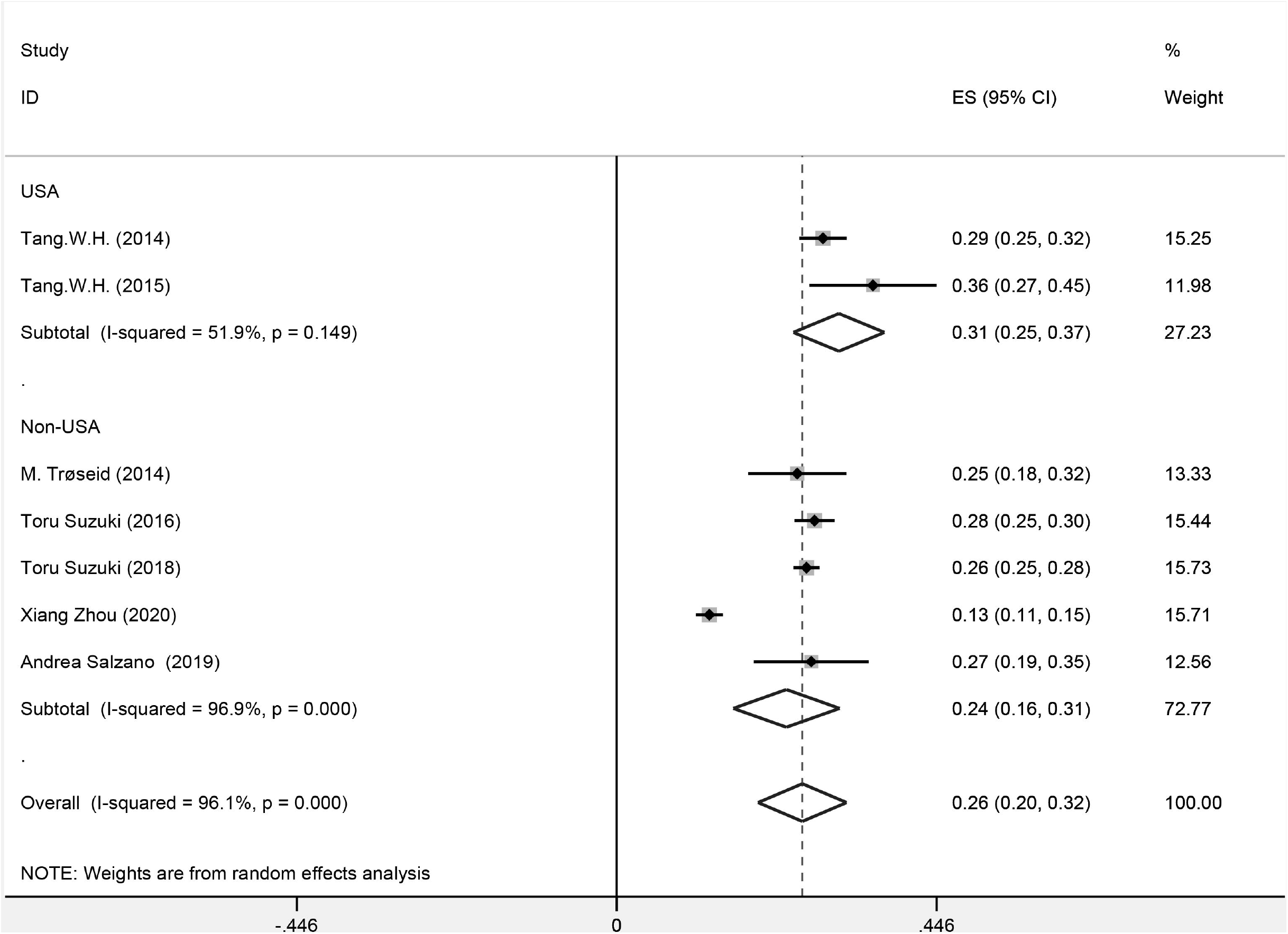
Forest plot for the association between TMAO and All‐cause mortality in the United States and those in other countries.

**Figure 11.**
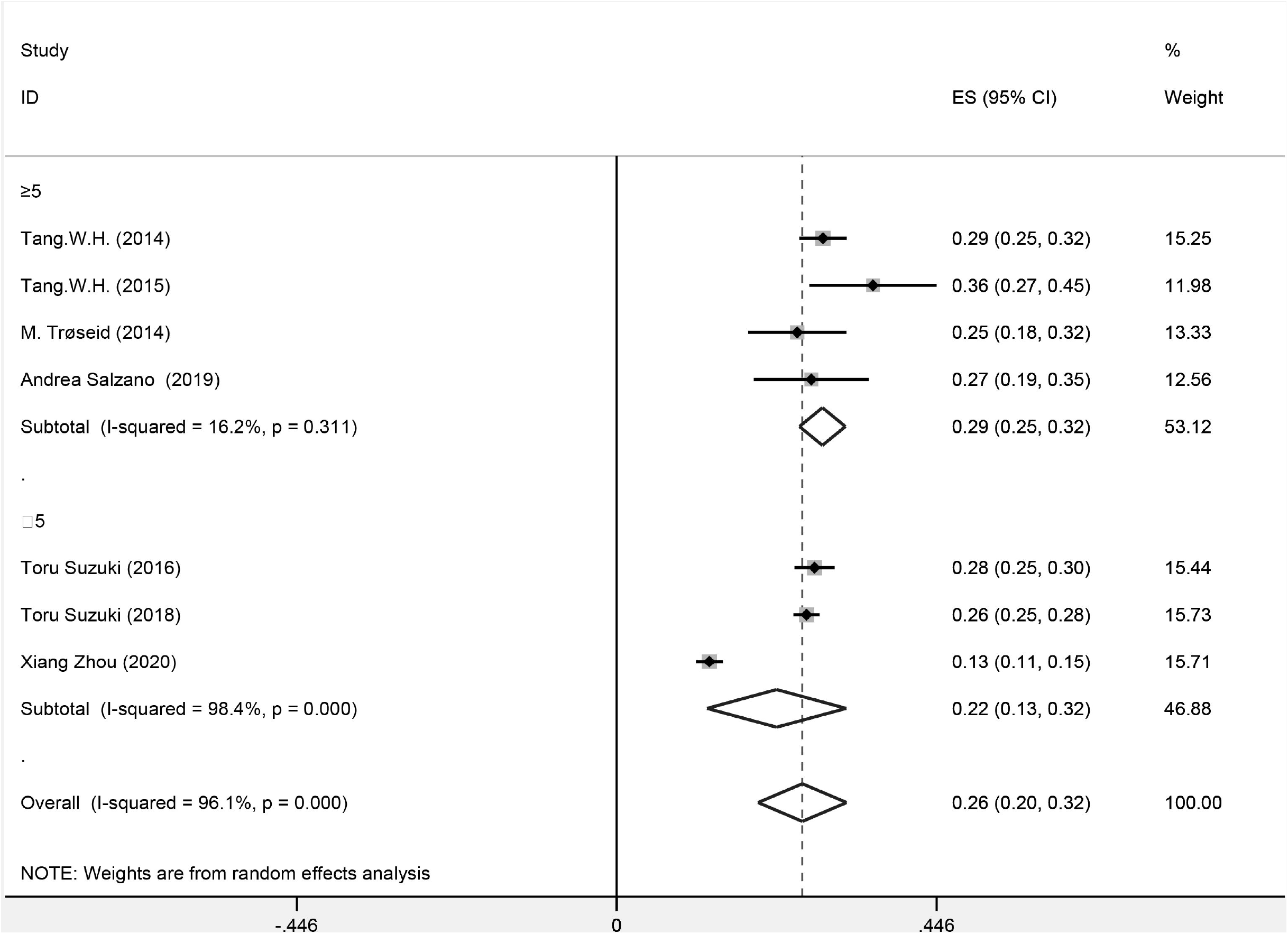
Forest plot for the association between TMAO and All‐cause mortality with different follow-up periods.

**Figure 12.**
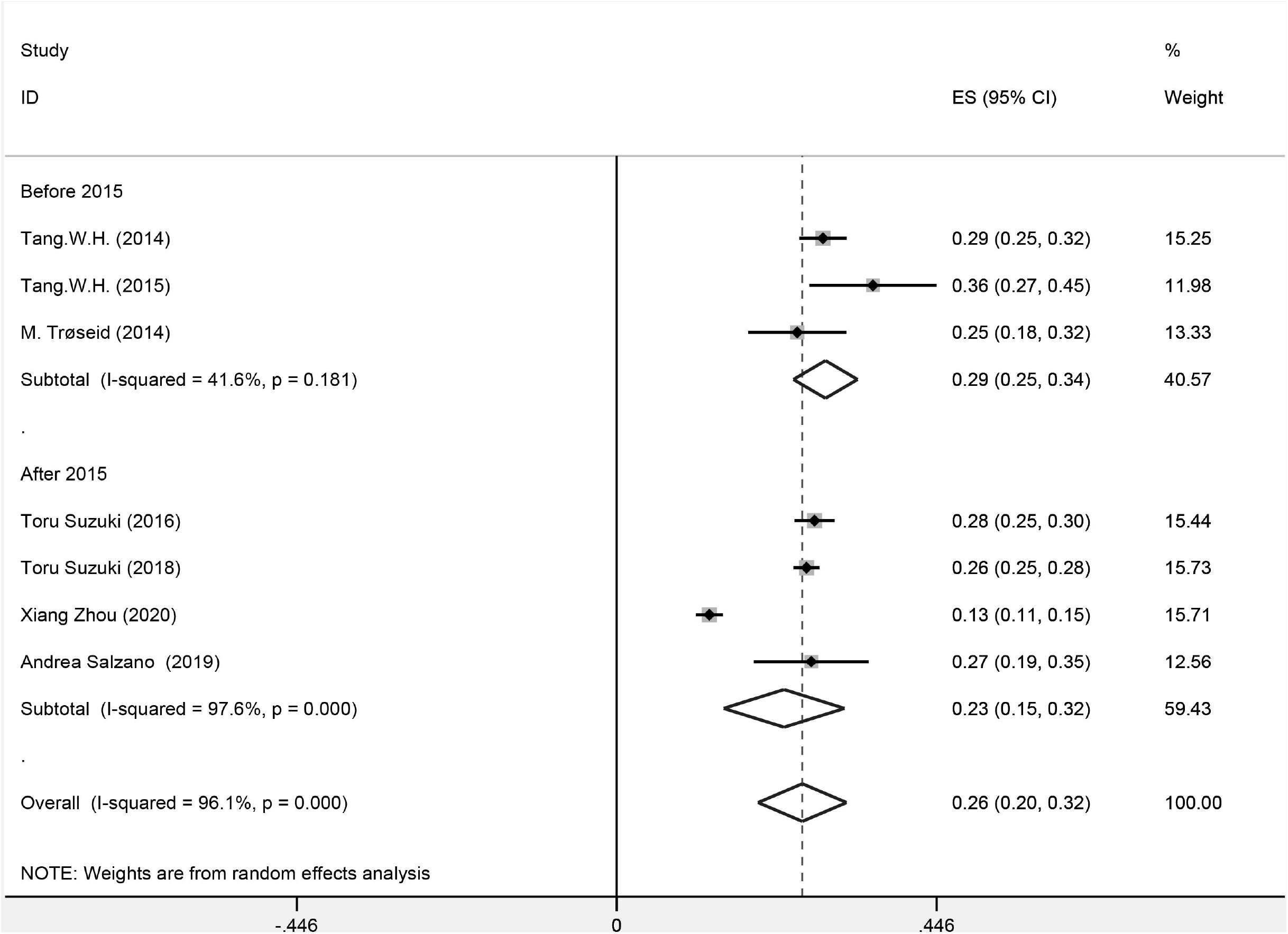
Forest plot for the association between TMAO and All‐cause mortality in studies before 2015 and after 2015.

**Figure 13.**
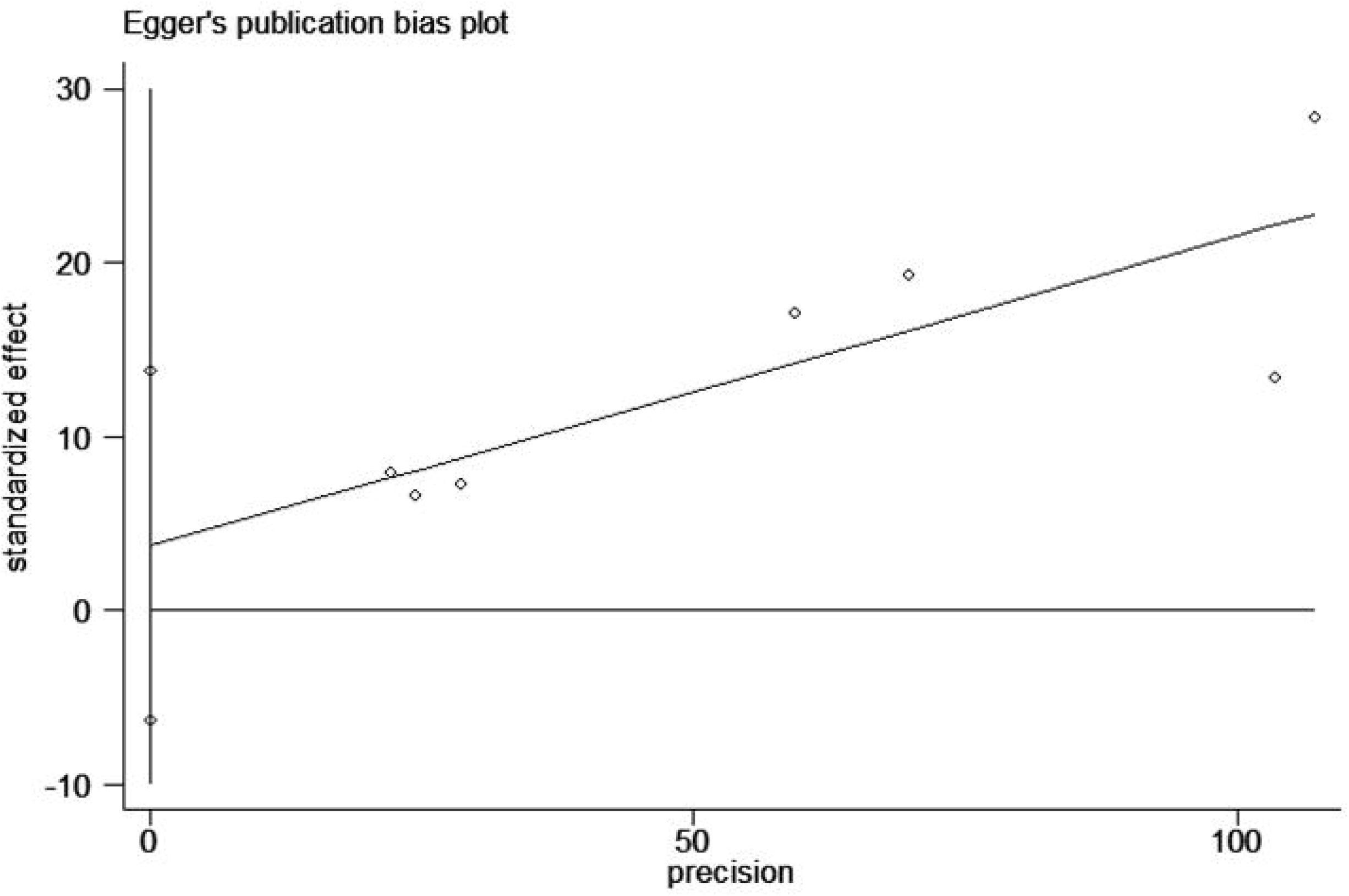
Egger’s publication bias plot for the association between baseline TMAO and All‐cause mortality.

**Figure 14.**
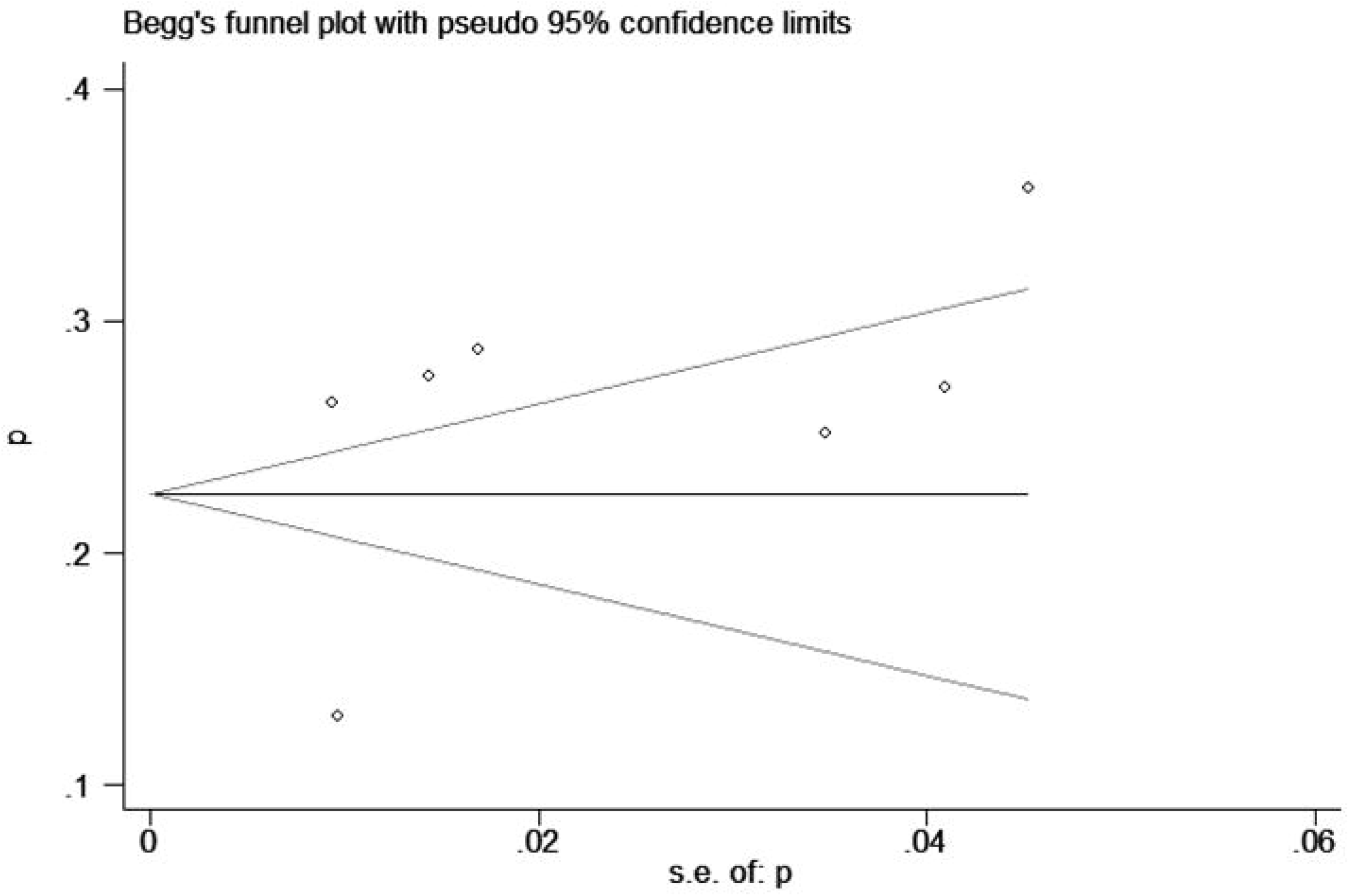
Begg’s publication bias plot for the association between baseline TMAO and All‐cause mortality.

**Figure 15.**
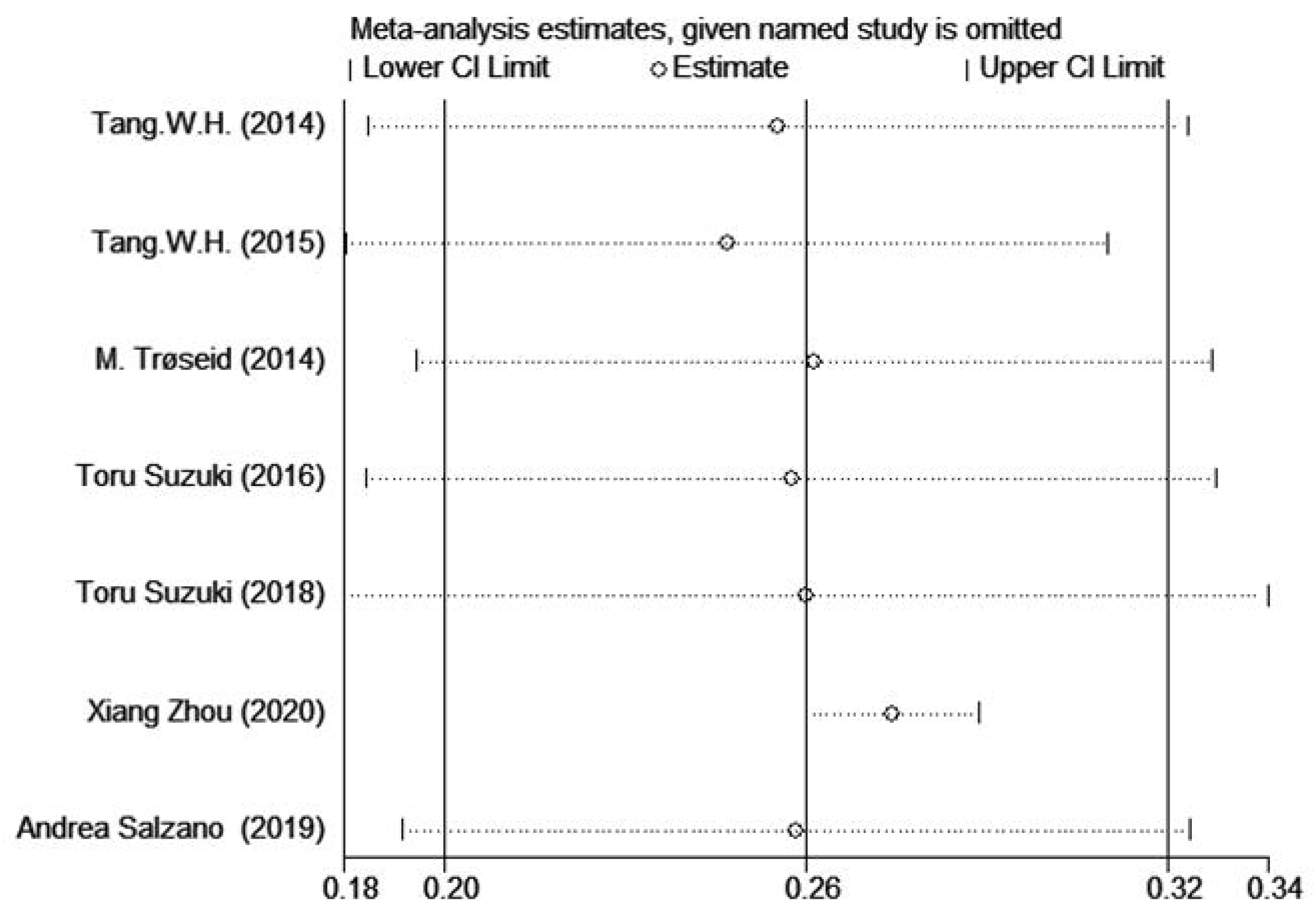
Sensitivity analysis for TMAO level and All‐cause mortality.

## Discussion

In this study, we concluded the baseline plasma TMAO was independently correlated with a 79% increased risk of adverse outcome, a 74% increase in all-cause mortality based on the data from 7 included publications with 5,519 sample size altogether. Although there was significant heterogeneity observed in correlationship between baseline TMAO and all-cause mortality and adverse outcomes, subsequent subgroups did not support follow-up time, study location, and year of publication characteristics significantly affected relationship between TMAO and adverse outcomes or all-cause mortality. These results indicate that subjects with a higher baseline level of TMAO are at greater risk of death from all cause and adverse outcomes in the future. High TMAO levels is an indication of poor prognosis of heart failure and can be used as an indicator to suggest further intervention for the patients in risk.

The mechanism of TMAO elevation in HF may be multifactorial. Choline is an essential nutrient for the synthesis of cell membrane phospholipids, methyl metabolism and the synthesis of neurotransmitter acetylcholine. Intestinal flora can convert phosphatidylcholine (lecithin) choline into trimethylamine (TMA), which is then absorbed by human hosts and converted into TMAO by liver flavin monooxygenase (FMO). The composition of intestinal flora is the main factor determining the level of TMAO. Intestinal flora composition deteriorated by HF can alter circulating TMAO levels. Excessive bacterial growth or changes in gut microbiota composition due to changes in microenvironment [14] or possible gut ischemia [15] in HF patients might be able to explain this development, since TMAO level can be changed by bacterial composition. The increase in dietary sources or microbial/host enzymes or the reduction in clearance due to renal insufficiency may also be a contributing effect. Since kidney is vital for TMAO clearance, the decrease of TMAO clearance rate may lead to the increase of TMAO in patients with impaired renal function. Similarly, plasma TMAO level was negatively correlated with eGFR. The level of TMAO in uremic patients was the highest. However, it can be significantly reduced by hemodialysis. It is clear that renal injury itself can lead to TMAO accumulation through reduced clearance, even though patients with renal function retention are likely to be in high TMAO levels. The mechanism of TMAO contributes to detrimental outcome in HF may be multifaceted.[16]. First, TMAO affects myocardial hypertrophy and fibrosis. In studies of choline-fed mice, elevated TMAO levels have a direct effect on myocardial fibrosis and the activation of related fibrogenic pathway due to the metabolism of the gut microbiota [17]. Li, Z., et al. found that TMAO promotes myocardial hypertrophy and fibrosis through Smad3 signaling pathway [18]. Subsequently, it was reported that 3,3-dimethyl-1-butanol (DMB) which is an inhibitor to TMA synthesis, could prevent myocardial hypertrophy and fibrosis. This discovery can also support the role of TMAO in ventricular remodeling [19]. Secondly, TMAO can induce inflammation. Seldin, M. M., et al. reported that TMAO can directly activate inflammatory pathways, such as NF KB signaling, a signal pathway discovered in inflammation of vascular smooth muscle cell in choline fed mice [20]. Thirdly, TMAO aggravates mitochondrial dysfunction. Makrecka-Kuka, M., et al. found that TMAO lead to the impairment of pyruvate and fatty acid oxidation in cardiac mitochondria [21]. These pathophysiological mechanisms may turn into a deteriorating loop, thereby further aggravating the process of HF. In addition, the correlation between TMAO and LV diastolic function index is stronger than that of systolic function, so TMAO may affect the slope of pressure volume fitting curve [8; 9]. However, the evidence of the link between TMAO and HF are still inconsistent with each other and insufficient. For example, seafood that is rich in TMAO has been reported to protect against cardiovascular disease rather than leading to a worse outcome. Therefore, we collected all current studies on the relationship between TMAO and the prognosis of heart failure for meta-analysis, with the purpose of determining the value of TMAO on the prognosis of HF.

Our results further confirm that a higher TMAO at baseline is linked with a higher risk of all-cause mortality in HF patients. TMAO may be a prognostic marker of HF. The results of this study also suggest that TMAO has an early warning effect on the prognosis of heart failure, which should be paid attention to by clinicians. In addition, the discovery of TMAO as a prognostic marker for HF has provided a possible way to handle HF by targeting gut microbiota and its metabolites. Based on this finding, it is reasonable to speculate that lowering the level of TMAO may be a new method to treat or improve the prognosis of HF. TMAO is a molecule produced by metabolism of choline, betaine and carnitine by intestinal microorganisms. Plasma TMAO level is determined by various factors which includes gut microflora, drug administration and hepatoflavin monooxygenase activity [22]. Many therapeutic strategies have been proposed to reduce TMAO levels, such as dietary regulation, the use of antibiotics, the application of microecological agents (prebiotics), inhibition of key mediators of metabolites, and transplantation of intestinal flora. Studies have shown that the use of broad-spectrum antibiotics such as ciprofloxacin and metronidazole can suppress TMAO levels almost completely [23].

Studies conducted in mice have shown that using a mixture of vancomycin, neomycin sulfate, metronidazole, and ampicillin can inhibit atherosclerosis caused by choline (a precursor of TMAO) in the diet [24]. Inhibition of plasma TMAO levels can inhibit the formation of macrophage foam cells [24]. Studies have shown that increasing choline in the diet leads to an increase in circulating TMAO levels, which has a negative effect on the structure and function of the heart during HF [25]. Intestinal flora transplantation has been used to treat a variety of gastrointestinal diseases, such as Clostridium difficile infection and inflammatory bowel disease, and can also be used to improve atherosclerosis [26]. At present, there is no research on whether reducing TMAO level can improve the prognosis of HF. Our research provides a theoretical basis for further research.

### Strengths and limitations

Our study concluded potential prognostic power of TMAO in HF populations. To the best of our knowledge, so far, no other meta-analysis was published prior to prove that high‐TMAO level is positively correlated with adverse events of HF. We used different statistical methods to identify the potential sources of heterogeneity. Meanwhile, there are also limitations that should be taken into account. First of all, the sample size and included studies were not large enough in our meta-analysis (7 studies enrolling about 5500 patients), which might lead to an inconclusive conclusion. However, we performed subgroup analysis and sensitivity analysis to prove that the results of the study are relatively robust. Secondly, TMAO values are defined according to the original text, and the original data cannot be obtained, which is graded by HR. Therefore, in the future, more and more studies will be conducted according to the specific values. Thirdly, the baseline clinical characteristics of each study include considerable heterogeneity including age, gender, and race. There may still be other potential factors that involved in the correlation of TMAO and HF adverse outcomes (e.g., related to diet history and / or physical exercise). In addition, uncontrolled factors such as dietary patterns and genetic variations may significantly affect TMAO concentrations. Additionally, although there is an analysis of influencing correction factors in single study, the correction factors are not uniform among the included studies, making it difficult to analyze the relationship between TMAO and the prognosis of heart failure after adjusting the correction factors (including renal function). Finally, the lack of continuous data makes it difficult to quantify TMAO differences between subjects with and without events.

## Conclusion

Higher baseline level of TMAO is associated with poor prognosis of heart failure,with an increased risk of all-cause mortality and adverse events.

## Data Availability

The raw data supporting the conclusions of this article will be made available by the authors, without undue reservation.

## Author Contributions

HQ and DS: conceptualization and supervision. LM and HQ: data curation, formal analysis, investigation, and methodology. JD and ZC: project administration. LM and HQ: software and writing (original draft). MG, JD and LM: writing (review and editing). LM and JD contributed equally to this manuscript. All authors contributed to the article and approved the submitted version.

## PubMed

((((((((((((((((((Heart Failure[Title/Abstract]) OR Cardiac Failure[Title/Abstract]) OR Heart Decompensation[Title/Abstract]) OR Decompensation, Heart[Title/Abstract]) OR Heart Failure, Right-Sided[Title/Abstract]) OR Heart Failure, Right Sided[Title/Abstract]) OR Right-Sided Heart Failure[Title/Abstract]) OR Right Sided Heart Failure[Title/Abstract]) OR Myocardial Failure[Title/Abstract]) OR Congestive Heart Failure[Title/Abstract]) OR Heart Failure, Congestive[Title/Abstract]) OR Heart Failure, Left-Sided[Title/Abstract]) OR Heart Failure, Left Sided[Title/Abstract]) OR Left-Sided Heart Failure[Title/Abstract]) OR Left Sided Heart Failure[Title/Abstract])) OR “Heart Failure”[Mesh])) AND ((((((((((((((((((((((2-Hydroxy-N,N,N-trimethylethanaminium[Title/Abstract]) OR Choline Citrate[Title/Abstract]) OR Citrate, Choline[Title/Abstract]) OR Choline O-Sulfate[Title/Abstract]) OR Choline O Sulfate[Title/Abstract]) OR O-Sulfate, Choline[Title/Abstract]) OR Choline Hydroxide[Title/Abstract]) OR Hydroxide, Choline[Title/Abstract]) OR Bursine[Title/Abstract]) OR Vidine[Title/Abstract]) OR Fagine[Title/Abstract]) OR Choline Bitartrate[Title/Abstract]) OR Bitartrate, Choline[Title/Abstract]) OR Choline Chloride[Title/Abstract]) OR Chloride, Choline[Title/Abstract])) OR “Choline”[Mesh])) OR (((((Trimethyloxamine[Title/Abstract]) OR trimethylammonium oxide[Title/Abstract]) OR trimethylamine N-oxide[Title/Abstract]) OR TMAO[Title/Abstract]) OR trimethylamine oxide[Title/Abstract])) OR beatine) OR TMA) OR L-cartine)

## Embase

#37’chloride, choline’:ab,ti 21

#36 ‘choline chloride’:ab,ti 1,638

#35 ‘bitartrate, choline’:ab,ti 1

#34 ‘choline bitartrate’:ab,ti 46

#33 ‘fagine’:ab,ti 0

#32 ‘vidine’:ab,ti 2

#31 ‘bursine’:ab,ti 12

#30 ‘hydroxide, choline’:ab,ti 0

#29 ‘choline hydroxide’:ab,ti 26

#28 ‘o-sulfate, choline’:ab,ti 0

#27 ‘choline o sulfate’:ab,ti 29

#26 ‘choline o-sulfate’:ab,ti 29

#25 ‘citrate, choline’:ab,ti 35

#24 ‘choline citrate’:ab,ti 41

#23 ‘2-hydroxy-n,n,n-trimethylethanaminium’:ab,ti 3

#22 ‘trimethyloxamine’:ab,ti 1

#21 ‘trimethylammonium oxide’:ab,ti 4

#20 ‘trimethylamine n-oxide’:ab,ti 1,926

#19 ‘tmao’:ab,ti 1,683

#18 ‘trimethylamine oxide’:ab,ti 303

#17#1 OR #2 OR #3 OR #4 OR #5 OR #6 OR #7 OR #8 OR #9 OR #10 OR #11 OR

#12 OR #13 OR #14 OR #15 OR #16 533,637

#16 ‘systolic dysfunction’:ab,ti 15,591

#15 ‘propofol infusion syndrome’:ab,ti 272

#14 ‘high output heart failure’:ab,ti 465

#13 ‘heart ventricle overload’:ab,ti 0

#12 ‘heart ventricle failure’:ab,ti 0

#11 ‘heart outflow tract obstruction’:ab,ti 0

#10 ‘heart arrest’:ab,ti 597

#9 ‘forward heart failure’:ab,ti 5

#8 ‘experimental heart failure’:ab,ti 608

#7 ‘diastolic dysfunction’:ab,ti 18,824

#6 ‘congestive heart failure’:ab,ti 53,445

#5 ‘cardiorenal syndrome’:ab,ti 1,376

#4 ‘cardiopulmonary insufficiency’:ab,ti 242

#3 ‘cardiogenic shock’:ab,ti 20,162

#2 ‘acute heart failure’:ab,ti 10,796

#1

## Cochrane Library databases

Search help View saved searches Save this search PrintView fewer lines

#1

MeSH descriptor: [Heart Failure] explode all trees MeSH

9176

#2

(Heart Failure):ti,ab,kw

(Word variations have been searched)S Limits

34586

#3

(Heart Decompensation):ti,ab,kw

(Word variations have been searched)S Limits

1526

#4

(Decompensation, Heart):ti,ab,kw

(Word variations have been searched)S Limits

1526

#5

(Heart Failure, Right-Sided):ti,ab,kw

(Word variations have been searched)S Limits

67

#6

(Heart Failure, Right Sided):ti,ab,kw

(Word variations have been searched)S Limits

515

#7

(Right-Sided Heart Failure):ti,ab,kw

(Word variations have been searched)S Limits

67

#8

(Right Sided Heart Failure):ti,ab,kw

(Word variations have been searched)S Limits

515

#9

(Myocardial Failure):ti,ab,kw

(Word variations have been searched)S Limits

8949

#10

(Congestive Heart Failure):ti,ab,kw

(Word variations have been searched)S Limits

6764

#11

(Heart Failure, Congestive):ti,ab,kw

(Word variations have been searched)S Limits

6764

#12

(Heart Failure, Congestive):ti,ab,kw

(Word variations have been searched)S Limits

6764

#13

(Heart Failure, CongestiveHeart Failure, Left Sided):ti,ab,kw (Word variations have been searched)S Limits

1

#14

(Left-Sided Heart Failure):ti,ab,kw

(Word variations have been searched)S Limits

73

#15

(Left Sided Heart Failure):ti,ab,kw

(Word variations have been searched)S Limits

1194

#16

#2 or #3 or #4 or #5 or #6 or #7 or #8 or #9 or #10 or #11 or #12 or #13 or #14 or #15 Limits

35698

#17

(Trimethyloxamine):ti,ab,kw

(Word variations have been searched)S Limits

4

#18

(trimethylammonium oxide):ti,ab,kw

(Word variations have been searched)S Limits

2

#19

(trimethylamine N-oxide):ti,ab,kw

(Word variations have been searched)S Limits

118

#20

(trimethylamine N-oxideTMAO):ti,ab,kw (Word variations have been searched)S Limits

0

#21

(trimethylamine N-oxideTMAO):ti,ab,kw

(Word variations have been searched)S Limits

0

#22

(Choline):ti,ab,kw

(Word variations have been searched)S Limits

1299

#23

(Choline Citrate):ti,ab,kw

(Word variations have been searched)S Limits

14

#24

(Citrate, Choline):ti,ab,kw

(Word variations have been searched)S Limits

14

#25

(Choline O-Sulfate):ti,ab,kw

(Word variations have been searched)S Limits

0

#26

(Choline O Sulfate):ti,ab,kw

(Word variations have been searched)S Limits

6

#27

(O-Sulfate, Choline):ti,ab,kw

(Word variations have been searched)S Limits

0

#28

(Choline Hydroxide):ti,ab,kw

(Word variations have been searched)S Limits

3

#29

(Hydroxide, Choline):ti,ab,kw

(Word variations have been searched)S Limits

3

#30

(Bursine):ti,ab,kw

(Word variations have been searched)S Limits

0

#31

(Vidine):ti,ab,kw

(Word variations have been searched)S Limits

0

#32

(VidineFagine):ti,ab,kw

(Word variations have been searched)S Limits

0

#33

(Choline Bitartrate):ti,ab,kw

(Word variations have been searched)S Limits

34

#34

(Bitartrate, Choline):ti,ab,kw

(Word variations have been searched)S Limits

34

#35

(Choline Chloride):ti,ab,kw

(Word variations have been searched)S Limits

59

#36

(Chloride, Choline):ti,ab,kw

(Word variations have been searched)S Limits

59

#37

#17 or #18 or #19 or #20 or #21 or #22 or #23 or #24 or #25 or #26 or #27 or #28 or #29 or #30 or #31 or #32 or #33 or #34 or #35 or #36 Limits

1369

#38

#16 and #37 Limits

10

#39

Type a search term or use the S or MeSH buttons to compose S MeSH Limits

N/A

Dear editor,

Attached please find our article entitled: Prognostic value of the level of gut microbe-generated metabolite trimethylamine-N-oxide in patients with heart failure:a meta-analysis and systematic review, which we would like to submit for publication in BMJ Open.

Heart failure (HF) is a complex multifactor illness that leads to enormous health and socio-economic burden. In vivo studies using animal models and humans suggested that intestinal microbiota likely played a big role in the development of a variety of diseases, including heart failure. Recent studies demonstrated that HF patients had significantly changes in both intestinal microbiota and intestinal permeability. Further research found that TMAO, a metabolite of intestinal flora, was likely connected to the disease emergence and progression of HF. Some studies have pointed out that TMAO level in patients with heart failure is relatively high compared with patients without and is related to the mortality and prognosis of heart failure. However, the relationship between TMAO and the outcomes of HF has not been evaluated. To investigate the relationship, systematic review and meta-analysis of all existing cohort publications to assess the relationship between plasma TMAO at baseline level and the prognostic outcome in HF patients.

## Recommended reviewer

1.Centre for Evidence-based medicine, Beijing University of Chinese medicine. Shi-Bing Liang.

2.Weijing FAN,MD Affiliations: Shuguang Hospital Affiliated to Shanghai University of Traditional Chinese Medicine Address: Shuguang Hospital Affiliated to Shanghai University of Traditional Chinese Medicine, No.528, Zhangheng Road, Pudong District, Shanghai City 201203, Shanghai, China.

Yours

Sincerely

Lina Miao, Ming Guo, Zhuhong Chen, Dazhuo Shi, Jianpeng Du

